# Respiratory Syncytial virus Strain Evolution and Mutations in Western Australia in the Context of Nirsevimab Prophylaxis

**DOI:** 10.1101/2025.05.07.25326957

**Authors:** Binit Lamichhane, Cara A Minney-Smith, Jake Gazeley, Ushma Wadia, David A Foley, Hannah C Moore, Jelena Maticevic, David W Smith, Paul Effler, Christopher C Blyth, David Speers, Avram Levy

## Abstract

**Background:** Nirsevimab is a long-acting monoclonal antibody used to prevent respiratory syncytial virus (RSV) infection in infants and high-risk children. During the 2024 RSV season in Western Australia, 23,525 doses were administered to infants entering their first season, and 1,233 doses to at-risk children. In this context, the selection and spread of escape variants is a potential concern. This study aimed to investigate nirsevimab-binding site mutations using both clinical and wastewater data.

**Methods:** We performed whole-genome sequencing on 382 clinical RSV samples and 12 wastewater samples collected between September 2023 and October 2024. RSV subtypes, genetic diversity, and mutations within the nirsevimab-binding region of the F protein were analysed. Phylogenetic analysis was conducted to assess lineage dynamics and the potential emergence of escape variants.

**Results:** RSV-A was the dominant subtype (61.8%), with RSV-B accounting for 38.2% of cases. No lineage shifts were observed following nirsevimab introduction and none of the known mutations associated with high-level nirsevimab resistance were detected in either clinical or wastewater samples. The prevalent RSV-B mutation combination (F:I206M:Q209R:S211N) was observed consistently but is not associated with reduced nirsevimab efficacy. Wastewater sampling, covering approximately 2 million people from Perth metropolitan region, confirmed findings from clinical sequences, reinforcing the absence of resistance mutations.

**Conclusion:** No evidence of nirsevimab escape mutations was found in either clinical or wastewater samples during the 2024 RSV season. Continued genomic surveillance, including wastewater monitoring, is essential to detect emerging resistance and ensure the long-term efficacy of prophylactic interventions.

## INTRODUCTION

Respiratory syncytial virus (RSV) is a major cause of acute lower respiratory tract infections, particularly among infants, older adults, and immunocompromised individuals [1, 2]. RSV is an enveloped, negative-sense, single-stranded RNA virus belonging to the *Pneumoviridae* family. Its genome encodes 11 proteins, including the attachment (G) and fusion (F) glycoproteins. RSV is categorised into two major subgroups, RSV-A and RSV-B, based on genetic and antigenic differences in the G protein [3, 4]. Globally, RSV is estimated to cause 33 million infections, three million hospitalisations, and over 100,000 deaths annually in children under five years of age, with the highest mortality rates observed in low- and middle-income countries [5]. RSV is also a major contributor to severe respiratory illness and hospitalisation in older adults and individuals with underlying conditions. Despite the burden RSV imposes on global healthcare systems, until recently, effective preventative measures have remained limited [6].

The monoclonal antibody, nirsevimab, has emerged as a promising prophylactic intervention, specifically designed to protect infants from severe RSV infections by targeting the prefusion F protein, a critical component in viral infectivity [7, 8]. The F protein is highly conserved, not associated with antibody-dependent enhancement, and serves as a primary target for therapeutic antibodies like nirsevimab [9]. Prior to nirsevimab, palivizumab was the only available monoclonal antibody for RSV prevention, but the need for monthly dosing restricted its widespread use [10]. RSV’s capacity for genetic variation, mainly driven by point mutations due to immune pressure, poses challenges for sustained efficacy of prophylactic and therapeutic interventions [11, 12]. The widespread introduction of nirsevimab may impose new selective pressures on circulating RSV strains [13]. Mutations within the nirsevimab binding region could potentially alter binding affinity, resulting in immune evasion and reduced prophylactic effectiveness, as seen with monoclonal antibody therapy for SARS-CoV-2, where changes in the spike protein made treatments less effective or even obsolete, especially for immunosuppressed patients [14]. Understanding how RSV strains evolve in the presence of targeted prophylaxis is crucial to monitoring its long-term effectiveness and potential impacts on viral dynamics.

Western Australia (WA) provides a unique opportunity to study RSV evolution in the context of widespread nirsevimab implementation, given its distinct RSV seasonality and extensive surveillance infrastructure [15]. In WA, where the majority of the state exhibits a temperate climate, seasonal RSV peaks are well-documented [16], and the introduction of nirsevimab offers a chance to assess its real-world impact on RSV strain evolution and mutation patterns. On 2 April 2024, the WA Department of Health launched the RSV Infant Immunisation Program where nirsevimab was offered to all infants born between 1 October 2023 and 30 September 2024 and all Aboriginal and/or Torres Strait Islander children and selected medically at-risk children entering their second RSV season born from 1 October 2022 to 30 September 2023. This introduction of nirsevimab offered an opportunity to assess its real-world impact on RSV strain evolution and mutation patterns. Overall nirsevimab coverage among the infant cohort in 2024 has been estimated to be 70% with 23,525 doses administered to infants entering their first season and 1,233 doses administered to at-risk children entering their second RSV season.

In this study, we investigated RSV strain evolution and mutations in WA during the first season of nirsevimab use. Whole genome sequencing was performed on a proportion of RSV detections to explore the prevailing subtypes and detect nucleotide changes within nirsevimab binding sites of the F-protein. We also assessed the suitability of wastewater surveillance for detecting community-level mutations. By integrating clinical and wastewater sequencing data, we aim to identify genetic changes in RSV, with a specific focus on mutations in the nirsevimab binding region. These findings will contribute to understanding the molecular epidemiology of RSV and the implications of nirsevimab on viral evolution in a real-world setting.

## METHODS

### Study population and specimen collection

Respiratory samples were selected to represent all age groups and collection periods from September 2023 to October 2024. These included all cases admitted to intensive care units (ICU) and breakthrough infections, spanning pre-, during, and post-peak RSV seasons. The non-breakthrough and non-ICU cases were randomly selected to represent different age groups and geographical regions. Breakthrough infections were defined as RSV detections in nirsevimab-eligible infants who had received nirsevimab. For wastewater surveillance, RSV-positive samples from July and August 2024 across metropolitan WA, identified as a period of elevated RSV concentration using real-time PCR, were chosen for whole-genome sequencing.

### Whole genome sequencing

Total nucleic acid was extracted from RSV-positive clinical respiratory specimens using MagNA Pure 96 DNA and Viral NA Large Volume Kit (Roche, Basel, Switzerland) using the manufacturer’s instructions. The extraction of wastewater specimens was performed according to established protocols [17]. cDNA was synthesised from the purified nucleic acid extract using LunaScript RT SuperMix Kit (New England BioLabs, Ipswich, MA, USA). The amplicons were generated using a tiled amplicon approach using ARTIC RSV A and B primer sets [18] in combination with Q5 Hot Start DNA Polymerase (New England BioLabs, Ipswich, MA, USA), following the manufacturer’s recommended protocol. The sequencing libraries were prepared using the Nextera XT DNA Library Preparation Kit (Illumina, San Diego, CA, USA) at a modified reagent volume ratio of 0.5. Genomic paired-end reads were generated using the Illumina MiniSeq short-read sequencing platform with the MiniSeq Mid Output Kit (300 cycles).

### Data analysis

The sequencing reads were quality-trimmed using BBDuk v37.99 and primers were removed with Cutadapt v1.18, followed by *de novo* assembly using MEGAHIT v1.2.9. The high coverage contigs (average *k*-mer coverage of at least 50) were blasted against an in-house reference database to determine the RSV subtype. Once the subtype was determined, trimmed reads were mapped to the corresponding reference genomes (NCBI Accession No: MH760627.1 for RSV-A, and MH760652.1 for RSV-B) using BBMap v37.99, and the consensus genome was extracted using ivar v1.2.2. Clade and genotype were assigned using Nextclade v3.8.2. Only samples that have greater than 70% coverage, 100X depth and nirsevimab binding site coverage were selected for further analysis.

The amino acid sequence of the F protein from the Nextclade output was imported into Geneious v2024.0.5 and analysed for amino acid substitutions at site φ, with the nirsevimab-binding site defined as residues 62-96 and 196-212.

Phylogenetic analysis was performed on 749 RSV-A and 916 RSV-B genomes, including previously sequenced cases from WA and sequences retrieved from GenBank and GISAID covering a range of sub-lineages and contemporary global genomes. Sequences were aligned using MAFFT v7.467 and the phylogenetic trees for RSV-A and RSV-B were constructed using RAxML v8.2.12 using the GTR-Γ nucleotide substitution model with 100 bootstrap replicates.

For wastewater samples, the primer and quality trimmed reads were mapped to reference F-protein genes of both RSV-A and RSV-B using BBMap v37.99. The resulting bam files were sorted and subsequently indexed using SAMtools v1.6 before variant calling with ivar v1.2.2.

## RESULTS

### Study population

From September 2023 to October 2024, there were 3,209 RSV detections with 41% (n=1312) of the patients between 1-4 years of age followed by 20% (n=637) younger than 12 months of age (Figure 1). Of these, 479 samples were sequenced in this study with 382 sequences passing quality control criteria for analysis. Of all the sequenced samples, 198 (51.7%) were from males, and 184 (48.3%) were from females with their ages ranging from less than one month to 92 years (median: 2 years). Samples from infants 12 months and younger accounted for a third of those analysed (32.4%, n=124), followed by persons aged 1-4 years (31.9%, n=122), 65-79 years old (10.2%, n=39), >80 years old (5.2%, n=20) and 5-15 years (5.2%, n=20) (Figure 1).

**Figure 1.**
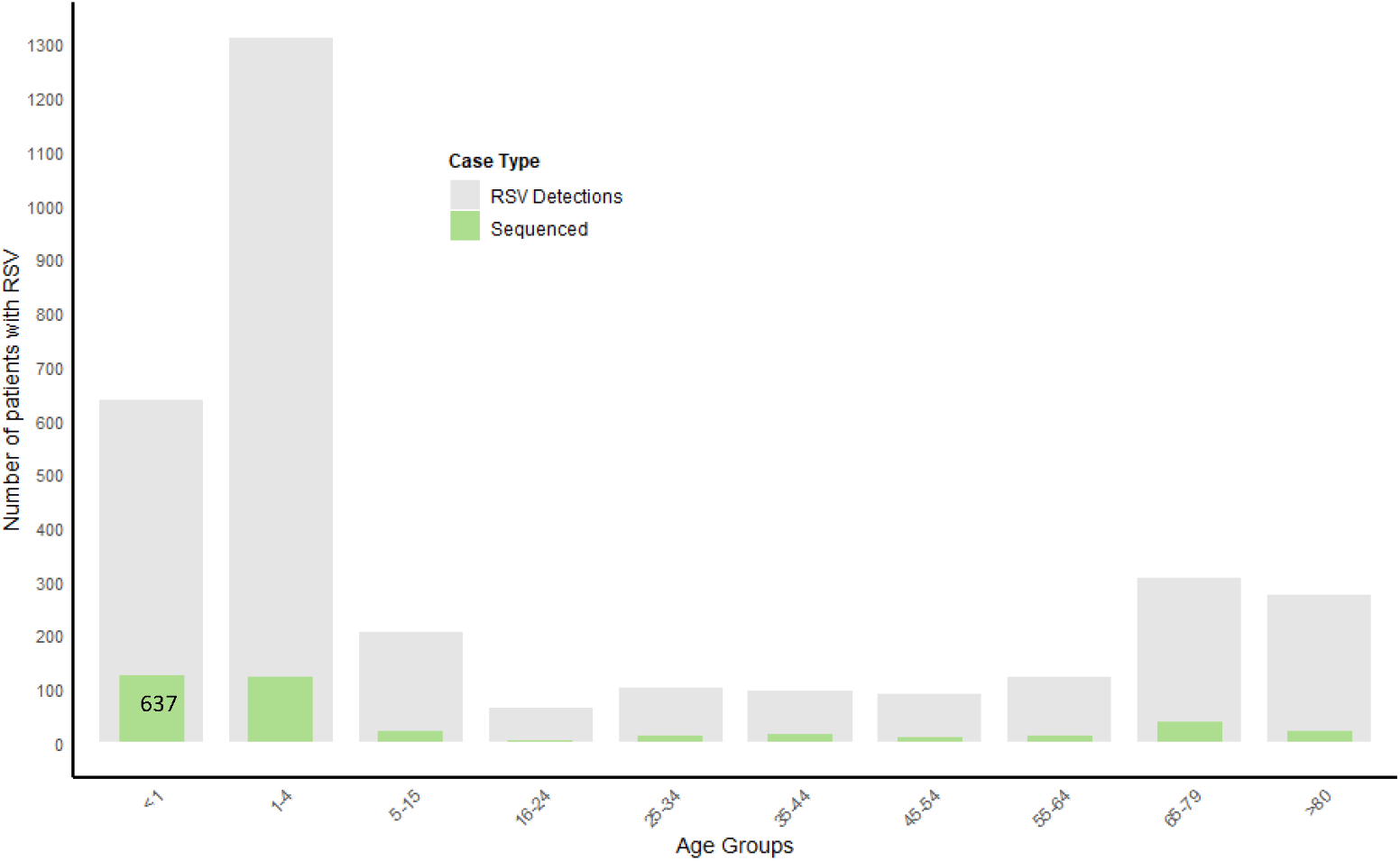
Total RSV detections and RSV samples sequenced from September 2023 to October 2024.

### Clinical RSV genotypes and lineages

Whole genome sequence analysis revealed that RSV-A was the most prevalent circulating subtype during the study period (236 [61.8%] of 382). All RSV-A samples belonged to the G.2.3.5 genotype and comprised nine RSV-A sub-lineages, all derived from lineage A.D., with the majority clustered in sub-lineages A.D.1.4 (n=74, 31.3%), A.D.3 (n=48, 20.3%), and A.D.1 (n=48, 20.3%) (Figure 2A). RSV-B subtype accounted for 146 samples (38.2%), with all belonging to genotype GB5.0.5a, including two sub-lineages circulating in 2024: B.D.4.1.1(n=40, 27.4%) and B.D.E.1 (n=105, 71.9%), both derived from lineage B.D. Notably, no B.D.E.4 cases were detected in 2024, despite its presence in 2023 (Figure 2B).

**Figure 2.**
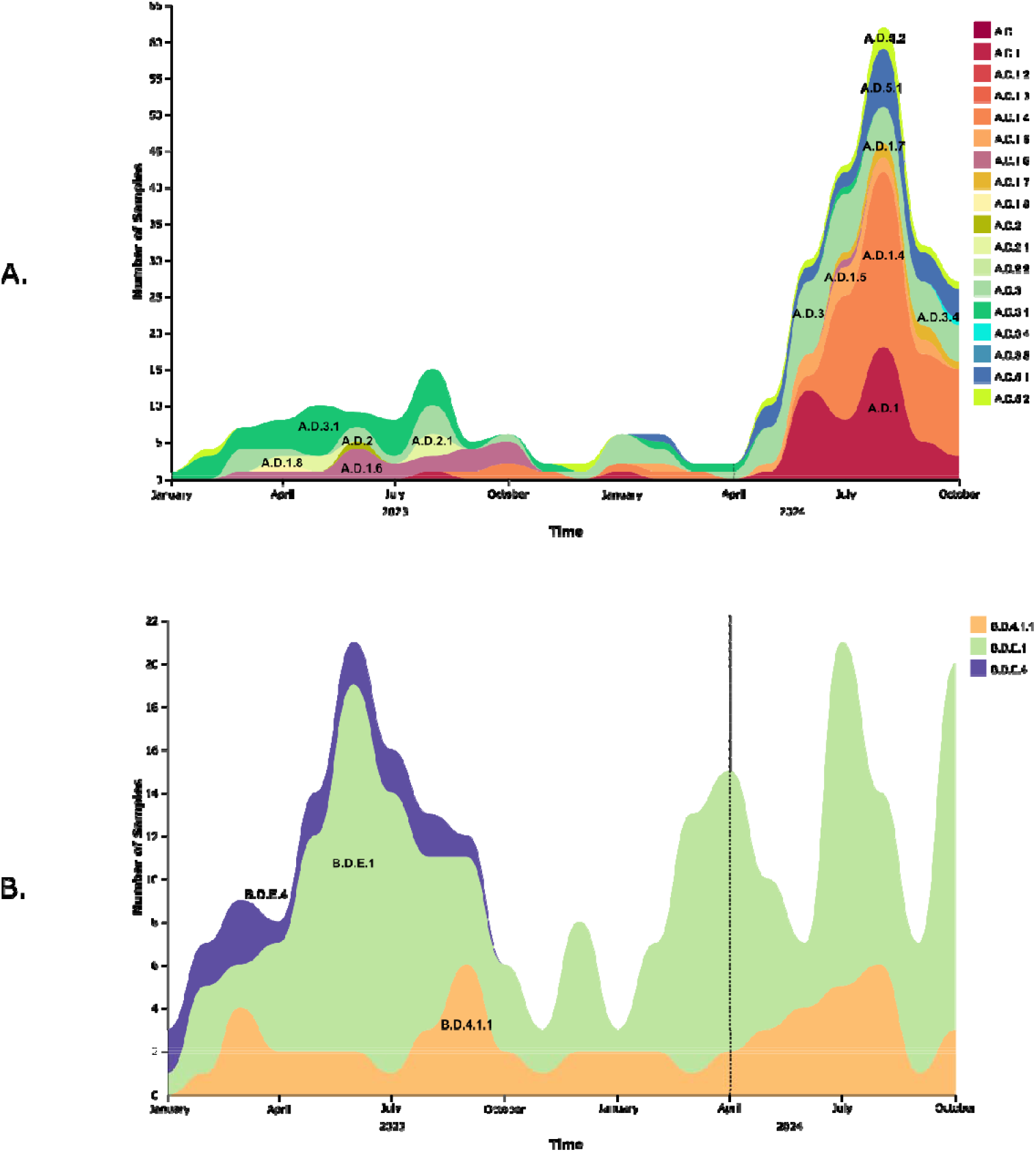
Circulating RSV-A (A) and RSV-B (B) lineages in WA before and during the study period. RSV samples collected between January and August 2023 are included as contextual references to provide a broad view of the lineages circulating prior to the nirsevimab rollout. The dashed line indicates the nirsevimab rollout date.

### Phylogenetic analysis

The phylogenetic analysis of RSV-A and RSV-B samples, compared with previous WA samples and global contextual data, revealed that the lineages circulating in WA were similar to those circulating globally, with no specific lineage or clade dominance observed both during and before the nirsevimab administration period (Figures 3 and 4). Furthermore, no age-specific phylogenetic clustering was observed. The predominant globally circulating strains in 2023–2024 are consistent with the RSV-A and RSV-B lineage data observed in WA (Figure 3-4).

**Figure 3.**
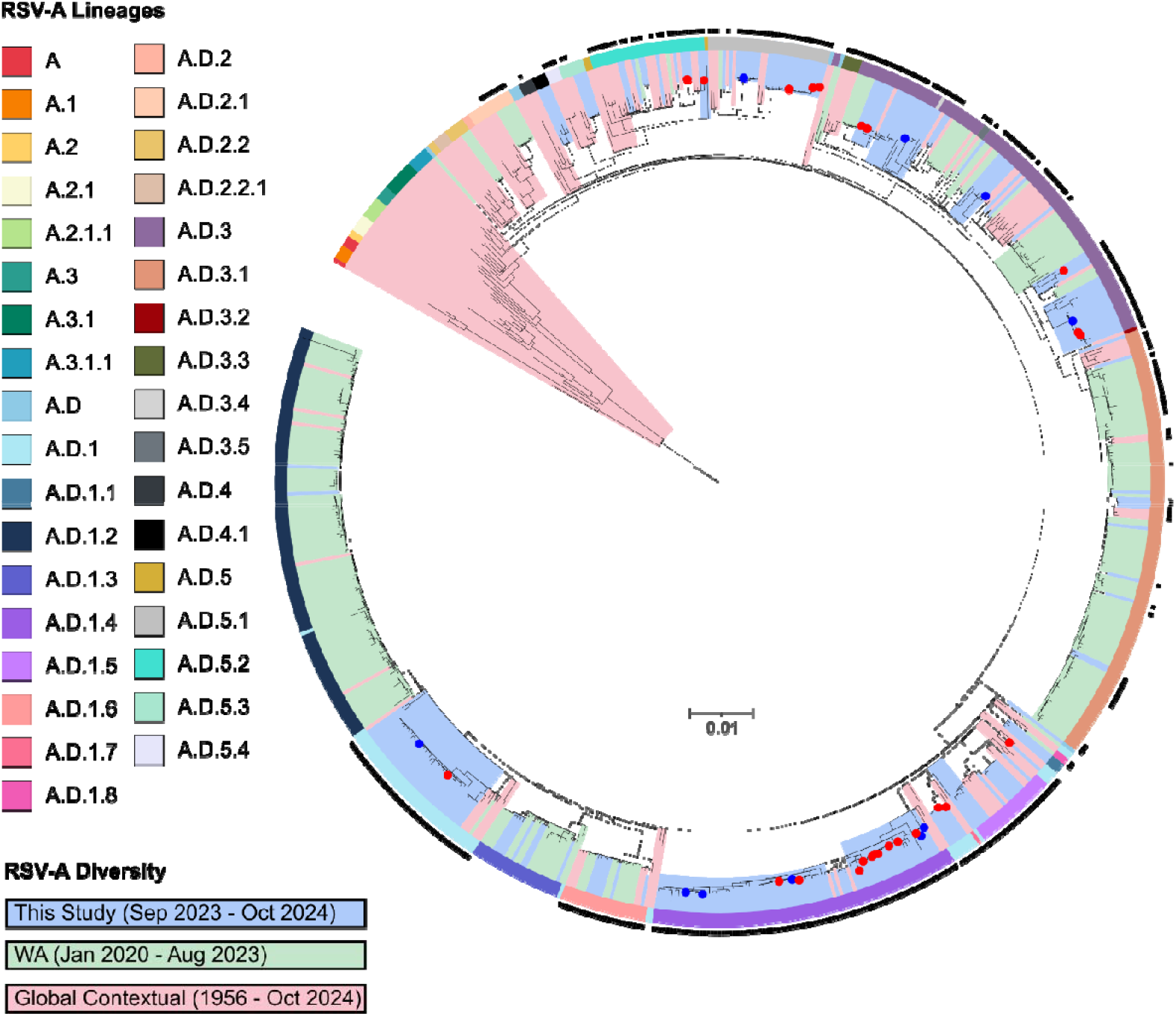
Maximum-Likelihood phylogenetic tree of clinical RSV-A, comprising 236 RSV-A samples from this study, 295 historical WA samples, and 218 global contextual samples sourced from GISAID and GenBank. Tip colours denote breakthrough cases (blue) and ICU cases (red). Contemporary sequences from 2023 and 2024, including global contextual samples, are highlighted with a black outer ring. RSV-A lineages are shown in the inner ring, coloured according to legend. The sequences from this study, previous WA sequences and globally-derived sequences are represented by clade colour according to the legend.

**Figure 4.**
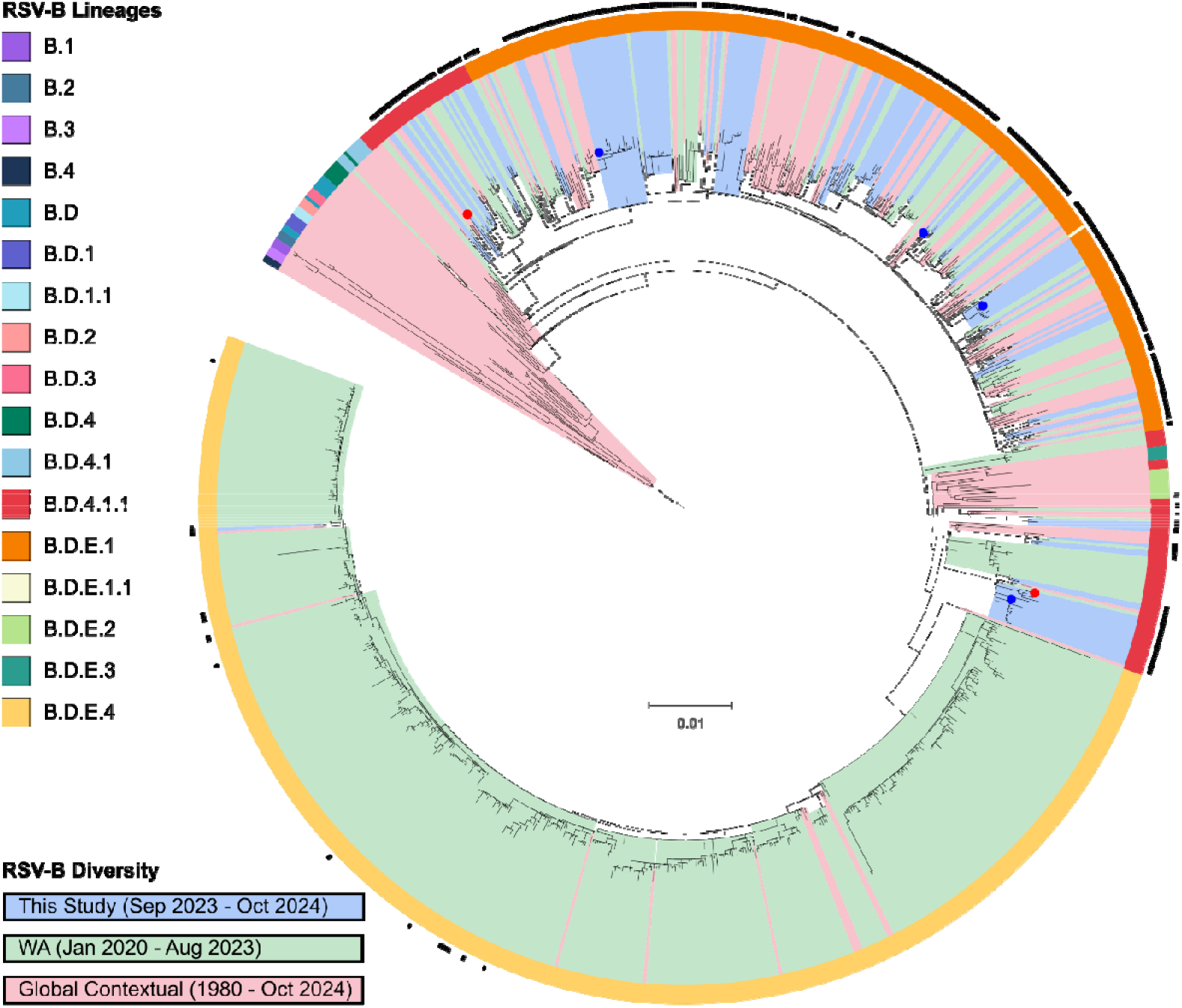
Maximum-Likelihood phylogenetic tree of RSV-B, comprising 146 RSV-B samples from this study, 578 historical WA samples, and 192 global contextual samples sourced from GISAID and GenBank. Tip colours denote breakthrough cases (blue) and ICU cases (red). Contemporary sequences from 2023 and 2024, including global contextual samples, are highlighted with a black outer ring. RSV-B lineages are shown in the inner ring, coloured according to legend. The sequences from this study, previous WA sequences and globally-derived sequences are represented by clade colour according to the legend.

### Breakthrough and ICU cases

Of the 382 samples sequenced, 26 (6.8%) were breakthrough infections and 16 (4.2%) cases were admitted to ICU including two breakthrough cases. Breakthrough cases occurred a median of 68 days post-Nirsevimab (range 13 to 140 days). Breakthrough cases were dominated by RSV-A (n=24, 10.2 % of total RSV-A cases) with only two RSV-B cases (1.4% of total RSV-B cases). Both ICU admitted breakthrough cases were RSV-A. A similar pattern was seen in ICU cases where 75% (n=12) were RSV-A (5.1% of total RSV-A cases) while 25% (n=4) were RSV-B (2.7% of total RSV-B cases). While breakthrough and ICU cases were observed more frequently in RSV-A than in RSV-B, the difference is not statistically significant, and the sample size is limited. The RSV-A breakthrough and ICU samples were not found to be associated with a particular sub-lineage but rather included multiple sub-lineages (Figure 3). Both RSV-B breakthrough cases were lineage B.D.4.1.1, as was one of the ICU cases, while the other three ICU cases were from lineage B.D.E.1 (Figure 4).

### Aminoacid variation in nirsevimab-binding site

Overall, the genetic diversity of RSV F protein remained low in RSV-A and RSV-B circulating during the study period. Among the 236 RSV-A F protein sequences, amino acid variations were observed in at least one sample at 41 (7.1%) positions including two substitutions at the nirsevimab binding site. This included F:N67S in one sample and F:D200N in two separate samples. All three samples were collected during the pre-nirsevimab period between November 2023 to February 2024, and the significance of these mutations on nirsevimab neutralisation is unknown. In RSV-B, 12 (2.1%) variable positions were observed. The majority of RSV-B samples carried the mutation combination F:I206M:Q209R:S211N (94.5%) including the breakthrough and ICU cases. This mutation combination has been reported as common among circulating B viruses and has not been associated with nirsevimab neutralisation [19-21].

### Wastewater samples

Twelve wastewater samples were sequenced and analysed for mutations in the F protein. While RSV subtypes could not be differentiated in wastewater samples because of low genome coverage, amino acid substitutions in the nirsevimab binding region were specifically assessed. F protein coverage ranged from 29.5% to 86.8%, with half of samples successfully covering the nirsevimab binding sites. No mutations apart from the prevalent RSV-B mutation combination (F:I206M:Q209R:S211N) were detected, which was present in all six samples.

## DISCUSSION

This study investigated RSV genomic diversity in WA during the 2024 season amid the implementation of a universal nirsevimab infant immunisation program. A total of 382 clinical genomes and 12 wastewater-derived composite sequences were analysed for circulating subtypes, and nirsevimab binding site genetic mutations. Most clinical detections were in young children, aligning with accepted RSV disease burden patterns [5]. Sequencing across age groups, including ICU and breakthrough cases, provided insights into RSV dynamics and demonstrated the stability of RSV in the setting of targeted prophylaxis, even in breakthrough cases.

The introduction of nirsevimab within the RSV infant immunisation program marks a significant step forward in RSV prevention as it has been shown to protect infants from severe RSV disease [7, 22, 23]. However, as a monoclonal antibody targeting a specific region of the prefusion F protein, nirsevimab may exert immune evasive selective pressure on circulating RSV strains [24]. We found that the RSV F protein exhibited low overall genetic diversity. While no mutations directly impacting nirsevimab binding [8, 20, 21] were identified in this study, the prevalence of RSV-A mutations F:N67S and F:D200N, observed prior to nirsevimab implementation warrants further investigation. Specifically, it is crucial to determine the potential impact of these mutations on nirsevimab neutralisation. F:N67S mutation has been previously reported in RSV-A samples in China [25], and F:D200N mutation has been reported in 0.03% of RSV-A samples in a large observational study by Wilkins and colleagues covering 17 countries [21]. However, no data is available about their effect on sensitivity to nirsevimab. RSV-B samples exhibited lower genetic variability, with 94.5% carrying the F:I206M:Q209R:S211N mutation combination, which has been commonly reported in recent RSV-B lineages [20, 21, 26] and is not associated with reduced nirsevimab efficacy [21]. Several amino acid substitutions associated with high-level resistance to nirsevimab have been identified by other studies, predominantly in RSV-B compared to RSV-A. For RSV-A, the F:K68E substitution has been implicated in reduced virus neutralisation by nirsevimab [8, 21]. In RSV-B, key resistance-associated mutations include F:I64M/K65E, F:K68N/E/Q, F:N201S/T, and N208D, which have been strongly linked to increased resistance to nirsevimab [20, 21]. Importantly, none of these resistance-associated mutations were detected in the samples analysed during this study. These findings are reassuring; however, they underscore the need for continued genomic surveillance to detect emerging resistance mutations that could compromise the effectiveness of nirsevimab over time.

The predominance of RSV-A over RSV-B during the 2024 season aligns with observations from other contemporary studies, suggesting a consistent pattern in RSV subtype distribution [20, 27]. Notably, no lineage shifts were detected in our study for either RSV-A or RSV-B following the introduction of nirsevimab, indicating that its introduction did not immediately have an impact on the effect in genetic composition of circulating strains. The absence of the RSV-B lineage B.D.E.4 in 2024, despite its presence in 2023, underscores the dynamic nature of RSV evolution. Phylogenetic analysis further revealed that WA RSV lineages closely resemble globally circulating strains, with no evidence of significant regional divergence during the study period. These findings highlight the importance of continued surveillance to monitor potential shifts in RSV population structure under selective pressures. As high-level resistance mutations to nirsevimab have been identified predominantly in RSV-B compared to RSV-A it is important this analysis is repeated in an RSV-B dominant season.

Wastewater-based surveillance has emerged as a complementary approach to clinical sampling, offering a non-invasive method to monitor population-level RSV diversity [28]. Despite challenges such as low genome coverage and inability to differentiate subtypes, the approach identified the prevalent RSV-B mutation combination (F:I206M:Q209R:S211N) in all six samples that covered the nirsevimab binding region. Wastewater sampling covers approximately 2 million people across the Perth metropolitan area, providing a far broader community-level representation compared to clinical sequences. If nirsevimab resistance were to emerge, it would likely first occur in breakthrough cases. The absence of resistance mutations in both breakthrough cases and the broader population assessed through wastewater analysis reinforces the reliability of these approaches for early detection and ongoing surveillance. The wastewater read-level results were consistent with the clinical case sequencing, yet were generated from far fewer samples, offering a complimentary and cost-effective approach for tracking community-wide lineage trends and detection of emerging variants. As wastewater concentration, amplification and analysis methods are refined, sensitivity and depth of information generated will improve.

Our study has several limitations. In particular, the wastewater samples were few, with limited data generated. The clinical samples analysed may not fully represent the general population of RSV-infected individuals in WA, as they were predominantly collected from hospital settings rather than primary healthcare facilities or the community. Additionally, the study’s focus on the F protein may overlook mutations in other genomic regions that could influence RSV’s transmissibility or pathogenicity.

As RSV interventions expand to include greater use of vaccines in both pregnant and older individuals, and interventions are used in combination (i.e. maternal vaccination and infant immunisation across Australia from 2025), understanding the interplay between viral evolution, transmissibility, and clinical severity will be critical. Future research should aim to expand genomic surveillance to include other regions of the RSV genome, investigate the long-term impact of nirsevimab on RSV evolution and its effectiveness against emerging variants and explore the integration of clinical, genomic, and environmental data to provide a holistic understanding of RSV dynamics.

## CONCLUSION

The molecular diversity of RSV variants during the 2024 season in WA was investigated in the context of nirsevimab’s introduction and widespread use. This first year of nirsevimab implementation offered a valuable opportunity to assess RSV evolution under targeted prophylaxis. Analysis of the nirsevimab-binding site yielded consistent and reassuring findings, with no evidence of emerging resistance, even among breakthrough cases at this stage. As more RSV prevention products are utilised in the community, continued genomic surveillance, complemented by innovative approaches like wastewater monitoring, will be crucial for tracking RSV evolution and safeguarding the long-term effectiveness of prophylactic interventions.

## AUTHORS CONTRIBUTIONS

Conceptualisation: AL; Methodology: BL, JG; Formal analysis: BL; Investigation: BL, CMS, JG; Resources: JM, DWS, DS, PE, CCB, AL; Writing – Original Draft: BL; Writing – Review and Editing: CMS, JG, UW, DAF, HCM, JM, DWS, DS, PE, CCB, AL; Visualisation: BL; Supervision: UW,DAF, HCM, JM, DWS, DS, PE, CCB, AL; Funding acquisition: PE, CCB, DS, AL. All authors have read and agreed to the published version of the manuscript.

## CONFLICT OF INTERESTS

The authors declare that there are no conflicts of interest.

## FUNDING INFORMATION

CCB is supported by an NHMRC Investigator Grant (APP1173163) and HCM is supported by a Stan Perron Foundation. The REVIVE study is supported by the Perth Children’s Hospital Foundation and the STAMP RSV program is supported by the Stan Perron Charitable Foundation.

## DATA AVAILABILITY

The RSV sequences used in this study will be made publicy available with publication.

## ETHICAL APPROVAL

This study was carried out under REVIVE (<u>RE</u>spiratory syncytial <u>V</u>irus Immunisation program - e<u>V</u>aluating <u>E</u>ffectiveness and impact) protocol which was approved by Child and Adolescent Health Service Human Research Ethics Committee (Research Governance Service 6789) and Ramsay Health Care WA/SA Human Research Ethics Committee (2024/ETH/0027).

## ACKNOWLEDGEMENTS

The authors would like to thank Ms. Catherine Pienaar and Ms. Joanne Harvey for their valuable assistance in collating patient information, Ms. Hannah Simpson for creating Figure 1, and Mr. Alex Xiao for doing most of the specimen retrieval and extractions. We would also like to thank PathWest Molecular Diagnostic Laboratory for providing access to testing results and samples and the PathWest Environmental Microbiology Unit for providing wastewater extracts as a part of WA wastewater surveillance program. We extend our gratitude to all the staff members of the Pathogen Genomics and Surveillance Unit at PathWest for their support throughout this work. In addition, we would also like to acknowledge STAMP RSV program and its investigators.

## Notes

### Competing Interest Statement

The authors have declared no competing interest.

### Author Declarations

This study was carried out under REVIVE (REspiratory syncytial Virus Immunisation program - eValuating Effectiveness and impact) protocol which was approved by Child and Adolescent Health Service Human Research Ethics Committee (Research Governance Service 6789) and Ramsay Health Care WA/SA Human Research Ethics Committee (2024/ETH/0027).

